# Association between antidepressants and antimicrobial resistance: a scoping review

**DOI:** 10.1101/2025.07.02.25330745

**Authors:** Bukola G Adekanmi, Charis A Marwick, Huan Wang, Karen N Barnett

**Author notes:** Corresponding Author: Dr Karen Barnett. Author Biography Statements. Bukola G Adekanmi (MPH) is a graduate of BSc Microbiology and Master of Public Health. Charis A Marwick (PhD) is a clinical reader in the School of Medicine, University of Dundee and a consultant in infectious diseases and general medicine in NHS Tayside. Huan Wang (PhD) is a principal investigator with expertise in statistics and epidemiology in the School of Medicine, University of Dundee. Karen N Barnett*(PhD) is a lecturer in public health and epidemiology, and population health researcher at the School of Medicine, University of Dundee.

## Abstract

**Background:** Antimicrobial resistance and depression are leading causes of global morbidity and mortality. Depression is the most common mental health disorder, with antidepressant use doubling between 2000 and 2022. Infectious diseases are increasingly difficult to treat due to some antimicrobial agents becoming ineffective. Antibiotic use is known to drive resistance in clinical bacterial infections, and laboratory studies have found that antidepressants have antibiotic properties, affecting the microbiota and stimulating mechanisms that help certain bacteria resist antibiotics.

**Aim:** To map and evaluate existing literature on the association between antidepressants and antimicrobial resistance.

**Methods:** This scoping review followed the modified Arksey and O’Malley’s framework. Systematic searches of MEDLINE, PubMed, Scopus and grey literature were conducted. Studies were screened for eligibility, assessed for risk of bias using ToxRTool, and analysed using a narrative synthesis.

**Results:** Ten articles met the inclusion criteria, eight laboratory studies and two reviews. Studies reported that exposure of bacteria to antidepressants increased antibiotic resistance. There were no eligible human or animal microbiome, or population studies.

**Conclusion:** This scoping review highlights the need for microbiome, clinical, and population studies. Associations between antidepressants and antibiotic resistance in clinical infections have implications for real-world practice, impacting clinical guidelines and patient care.

**Key Messages:** **What is already known on this* topic:* Depression and antibiotic resistance are both critical global health concerns. The use of antidepressant drugs is increasing and there is a growing body of laboratory evidence demonstrating that antidepressants, particularly selective serotonin reuptake inhibitors (SSRIs), exhibit antimicrobial activity against bacteria.

*What this study adds:* This scoping review maps and synthesizes existing evidence on associations between antidepressants and antimicrobial resistance, including mechanisms involved, and identifies important gaps in the literature. The eight primary research studies included were all laboratory-based, bacterial studies. All found that exposure of bacteria to antidepressant drugs increased antibiotic resistance. There were no eligible human or animal microbiome studies, or human clinical or population studies.

*How this study might affect research, practice or policy:* This scoping review highlights the need for more microbiome, clinical, and population studies. Associations between antidepressants and antibiotic resistance in clinical infections could have implications for real-world practice, impacting clinical guidelines and patient care.

## INTRODUCTION

Depression is the leading mental health disorder globally, affecting people of all ages, gender and social background (1). The World health Organization (WHO) has projected that untreated mental health, particularly depression, will become the principal cause of global disease burden by 2030 (2). The use of antidepressant drugs is also increasing with reports of a doubling across Europe in the last two decades between 2000 and 2022 from 30.5 to 70.5 defined daily dose (DDD) per 1000 people (3, 4). Recent data shows that in both the UK and the USA, approximately one fifth of the population have reported symptoms of depressive disorder and have received at least one prescription for an antidepressant drug (5–7).

The ability of bacteria, viruses, fungi and protozoa to resist major antimicrobials used against them in infections in humans is widely recognised. The rise in antimicrobial resistance across the globe, has the potential to significantly impact population health as infections become harder to treat and clinical procedures such as surgeries become much riskier (8). There is established evidence from clinical and epidemiological studies that antibiotic use is a key driver of antibiotic resistance in clinical infections in individual people and at national population levels (9, 10). Misuse, for example, inappropriate prescribing and/or failure to complete a course of antibiotics as directed, contributes to antimicrobial resistance (11). Depression is associated with an increased risk of being hospitalized for a range of infections (12) and, several studies have reported that antidepressants have antimicrobial properties that contribute to the alteration of microorganism’s structure, diversity and numbers.

Laboratory studies have demonstrated the extensive impact of non-antibiotic drugs on gut microbiome composition, with one study identifying common resistance mechanisms conferred by antibiotics and other drugs prescribed in clinical practice including antidepressant drugs (13). In a recent review Guo et al discussed how antidepressants can help bacteria resist antibiotics. His laboratory study on *Escherichia coli* revealed that the microbiomes of mice treated with antidepressants were altered (14). This is significant because the gut microbiome plays a key role in various aspects of health, including metabolism, immune function, and even mental health.

In the face of the rising prevalence of depression and antidepressants prescribing, the link with antimicrobial resistance warrants further research. The aim of this scoping review is to: (i) map the existing evidence on antidepressants use and antimicrobial resistance including both human and laboratory studies, (ii) explore the association between antidepressants and antimicrobial resistance, and (iii) identify gaps in the existing evidence to inform future research areas.

## METHODS

The Arksey and O’Malley’s framework (15) modified by the Joanna Briggs Institute (JBI), and PRISMA for Scoping Reviews (PRISMA-ScR) (16) were applied. The key stages included: search strategy; source of evidence, screening and selection; data charting; quality appraisal; analysis and presentation of results.

### Framework

The Population, Concepts and Context (PCC) question framework (17) informed the search strategy. While for human studies the population of interest was people receiving antidepressant medication, population terms were not included because the review also considered laboratory-based studies. The Concept captured antidepressants and antimicrobial agents including antibacterial (antibiotics), antiviral, antifungal, and antiprotozoal drugs. The Context included all types of evidence (published and unpublished) from any country. The search strategy was designed using keywords and their synonyms as well as medical subject headings (MESH) for antidepressants and antimicrobial resistance. Boolean operators “OR” and “AND” were used to streamline, broaden, and combine the search. The final search strategy is detailed in Supplementary file 1.

### Source of Evidence

The main literature search used Medical Literature Analysis and Retrieval System Online (MEDLINE), PubMed and Scopus. Discovery Research Portal, Electronic thesis Online Service (EthOS) and Google Scholar were used to review the grey literature.

#### Screening and Selection

##### Eligibility criteria

Articles exploring antidepressants and antimicrobial agents (antibiotics, antifungal, antiviral, and/or antiprotozoal agents), and focussing on the relationship between antidepressants or antidepressant properties and antimicrobial resistance, were included. Articles written in English language and published between 1^s^ ^t^ January 2013 and 1^st^ April 2024 were included. All types of study design were considered including published peer-reviewed research and grey literature (See Supplementary Tables I & II).

##### Exclusion Criteria

Articles written in languages other than English were excluded. Articles reporting potential antimicrobial properties of antidepressants without reference to associations with antimicrobial resistance were excluded.

##### Study selection

After removal of duplicates, all citations retrieved during the electronic database searches were independently screened for eligibility by two reviewers (BA and KB). The same two reviewers independently screened all title/abstracts and full-text review. Any conflicts were resolved through discussion. The reference lists of all included articles were screened for any additional eligible studies, not already identified through the database searches.

### Data Charting

A data charting form, following the PRISMA-SCR (16) was used to extract the following information from each included article: author and year of publication, study type or design, aims/purpose, country/location, the number and type/class of antidepressants, antimicrobial drugs tested, test organism, methodology, and main findings.

#### Quality Assessment

The quality of the included papers was assessed using the Toxicological data Reliability Assessment Tool (ToxRTool) (18). ToxRTool was chosen because it provides comprehensive criteria and guidance for evaluating the inherent quality of studies focusing on toxicology data including substances (drugs). Each criterion on the ToxRTool reliability assessment was assigned 1 when the criterion was met and 0 when criterion was not met. Article that scored 15 to 18 (highest) were classified as reliable without restrictions, 11 to 14 as reliable with restrictions, and < 11 as unreliable (19).

### Analysis and Presentation of Results

Reporting followed the PRISMA and Meta-Analysis extension for Scoping Reviews (PRISMA-ScR) guideline (16). Heterogeneity in eligible studies preluded meta-analyses so a narrative synthesis is presented.

### Patient and Public Involvement

Patients or the public were not involved in the design, conduct, reporting or dissemination plans of our scoping review.

## RESULTS

The search strategy identified 520 articles, with 436 remaining after removal of duplicates (Figure 1). There were no relevant articles from Discovery Research Portal or EthOS. Title and abstract screening excluded 354 articles with 82 full text articles assessed. The most frequent reason for exclusion was that the focus wasn’t on the association between antidepressants and antimicrobial resistance (n = 46). See Supplementary Table III, for excluded studies following full-text review. Ten articles met the inclusion criteria. No additional studies were identified from the reference lists of the included studies.

**Figure 1:**
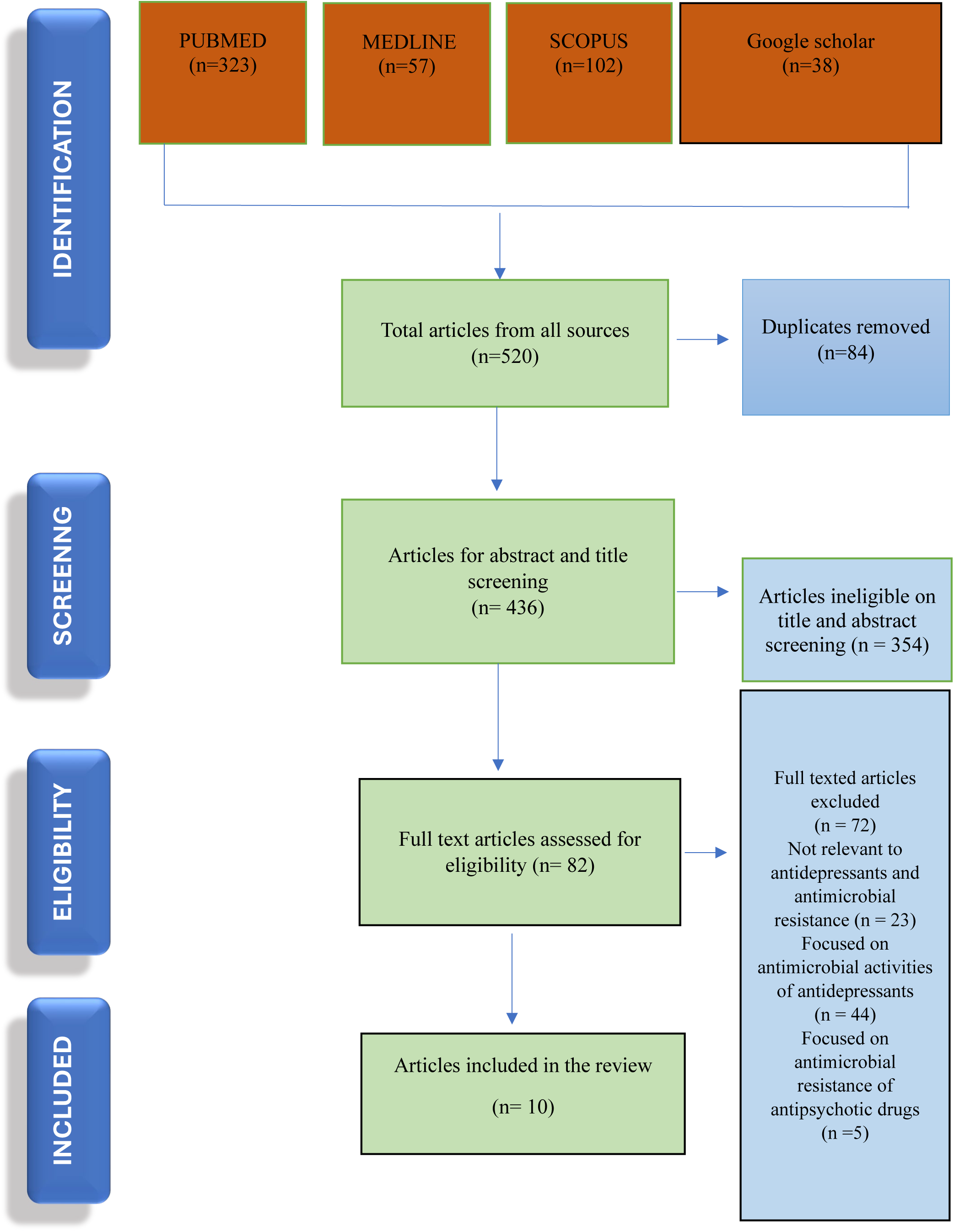
Prisma-ScR flow diagram of article identification, screening and selection.

### Characteristics of included studies

The 10 included studies were eight laboratory studies and two review articles (Table I). Although the search strategy was designed to identify both laboratory and human studies, there were no clinical, epidemiological, or population-based studies that were eligible for inclusion. Included articles were published between 2018 and 2023.

Geographically, studies originated from five academic institutions across five countries - Australia (three studies), Turkey (three studies), China (two studies), Canada (one study) and the USA (one study) (Table I).

### Toxicological data Reliability Assessment Tool (ToxRTool) Quality Assessment

Using the ToxRTool six studies were rated reliable without restriction (highest quality), (two were rated reliable with restrictions (medium quality) and two study was rated as not assignable due to lack/insufficient experimental details (19, 28)(See Supplementary Table IV).

### Pharmacological and Microbiological Characteristics

The included studies mainly focused either on the connection between antidepressants and antibiotics resistance in certain bacteria (n = 6) (13, 21, 23, 24, 27, 29) or evaluating the mechanisms employed by different antidepressants in promoting antibiotic resistance (n = 2) (20, 26). The study conducted by Wang et al (2023) (27) demonstrated that antidepressants can induce antibiotic resistance and persistence. Both reviewed articles (n =2) (14, 22) discussed the potential clinical implications of these findings. Seven primary studies (13, 20, 21, 23, 26, 27, 29) used a combination of phenotypic methods, including disc diffusion antibiotic susceptibility testing and minimum inhibition concentration (MIC) calculations, and genotypic methods, mainly polymerase chain reaction (PCR) and whole genome sequencing (WGS). One study (30) used mathematical modelling to predict antimicrobial resistance as an outcome of antidepressants exposure.

The organisms under investigation varied across the studies but all eight primary studies used Gram-negative bacilli - *E. coli*, *Actinobacter baylyi*, *Actinobacter baumannii*. Five studies used *E. coli* (four K12 (widely studied laboratory strain)(20, 23, 27, 29) and one wild type(13)), two studies used *A.baumannii* (21, 24) and one study *A.bayyl* (*26*).

A total of 15 antidepressant drugs from different classes, were tested for their potential to induce resistance to commonly used antibiotics across included studies (Figures 2&3). One study examined 12 (25), two studies each examined a different set of six (20, 26) one examined five (27), and two studies examined the same three (21,24) antidepressants. One (23) study focused on the underlying mechanism used by fluoxetine in activating multiple drug resistance and another one (13) on the synergistic effect of duloxetine and^2^chloramphenicol. The two review articles (14, 22) referenced primary laboratory studies (27) e^3^mphasizing the need for further research in establishing the association between antidepressa^5^nts and antimicrobial resistance, particularly through in-vivo experiments and population-based^1^studies.

**Figure 2:**
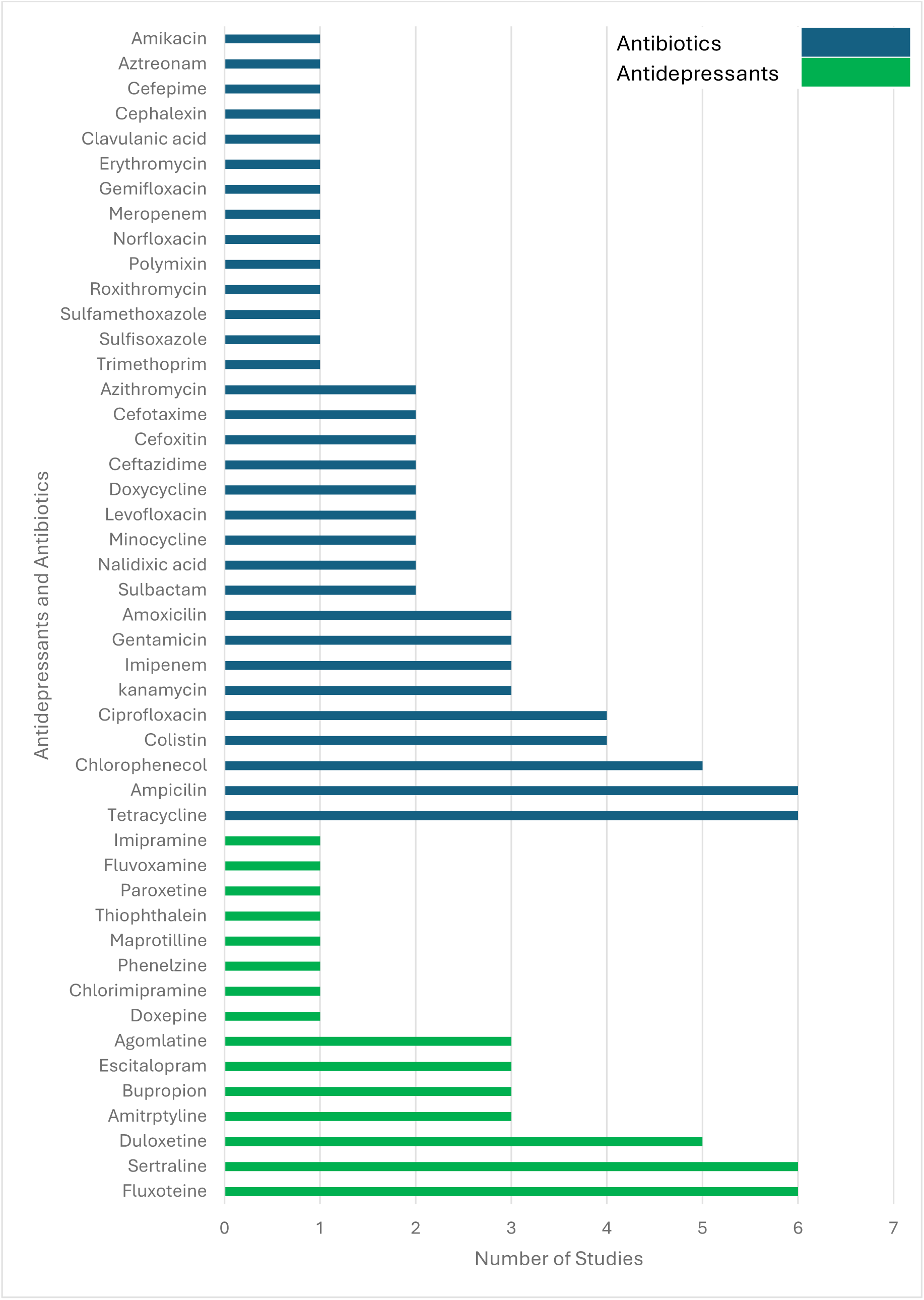
Antidepressants and antibiotics examined across the included studies.

**Figure 3:**
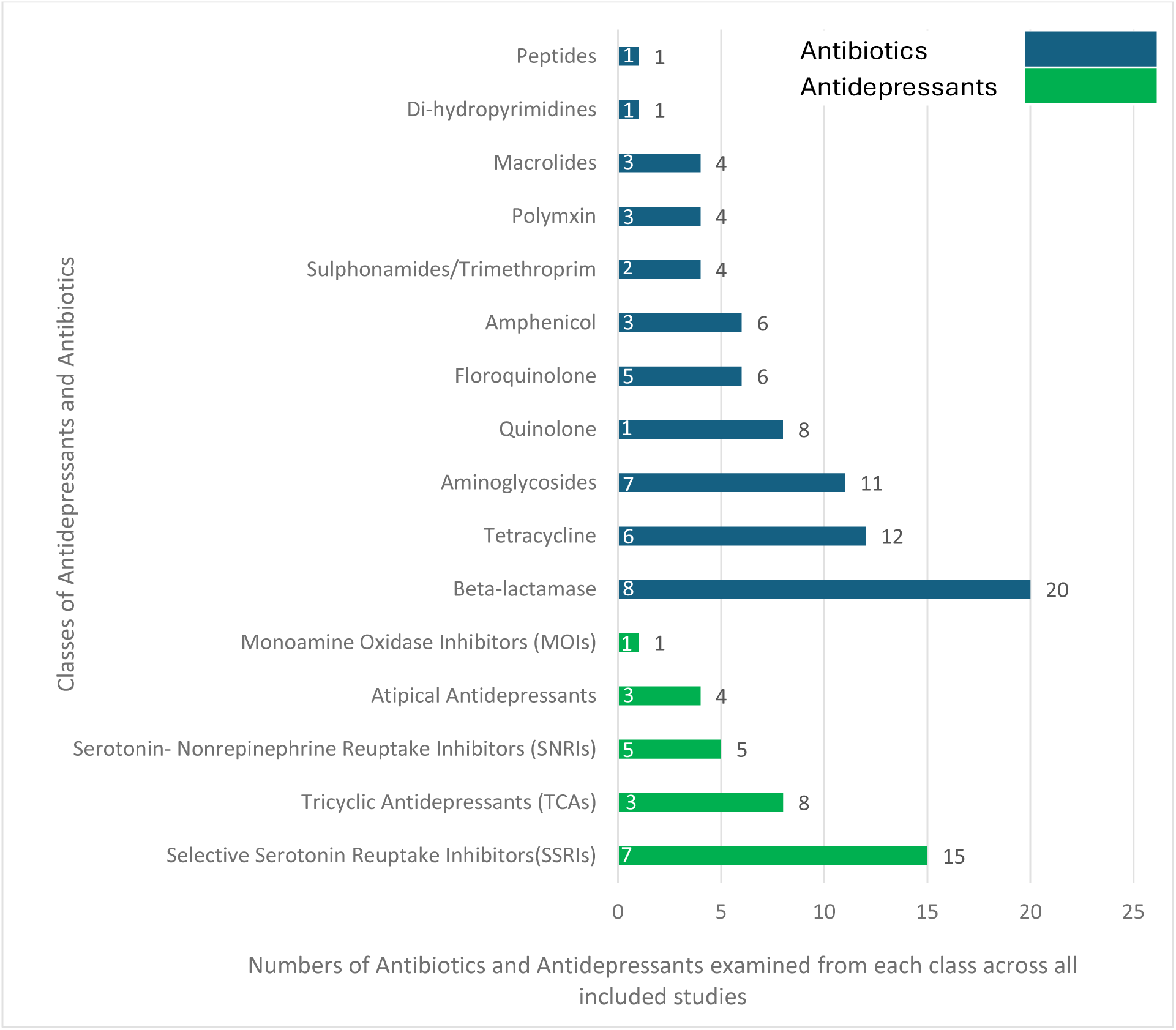
Classes of antidepressants and antibiotics examined in the included studies. Numbers in white represent the number of studies that included each class.

A total of 34 different antibiotics were used to evaluate resistance exhibited by organisms exposed to antidepressants (Figures 2&3). Beta-lactams were the most common class with 20 antibiotics tested across 6 studies (13, 20, 23, 26, 27, 29). Tetracycline and Aminoglycosides classes were also commonly tested with 12-and 11antibiotics included in six (13, 20, 23, 26, 27, 29) and seven studies (13, 20, 21, 23, 24, 26, 29) respectively. Of the eight experimental studies, seven focused on the resistance to several classes of antibiotics while one study focused on only two antibiotics from the following classes; Tetracycline and beta-lactamase.

### Mechanisms of Resistance Induced by Antidepressants

Six studies evaluated specific mechanisms of action of antidepressants in inducing antibiotic resistance (13, 20, 21, 23, 26, 27, 29). The role of oxidative stress in promoting fluoxetine-induced mutation in *E. coli* was examined by testing the susceptibility of the mutant strains against four different antibiotics (23). Antibiotic resistance in *E. coli* K12 was investigated by assessing reactive oxygen species (ROS) production and cell permeability through flow cytometry, proteomics, and RNA sequencing to analyze gene expression in response to antidepressant exposure (20). Phenotypic and genotypic analyses investigated links between ROS production and, resistance and persistence following antidepressant exposure in *E. coli* (27). The transcriptional response of environmentally isolated *E. coli* to antidepressant exposure was examined using both phenotypic and genotypic approaches (23). One study investigated the synergistic effects of antidepressants and antibiotics in stimulating resistance in *E. coli*, and analysed the mechanism underlying any collaborative enhancement (13). The expression of antibiotic resistance in *A. baumannii* was assessed by measuring efflux pump and outer membrane porin gene levels using quantitative reverse transcriptase PCR after 30 days of exposure to three different antidepressants (21). One study examined whether antidepressants can promote the changes and movement of antibiotics resistance genes in bacteria (26).

Two of the studies reported effects of antidepressants exposure on the mode of transfer of multiple resistance gene: horizontal conjugative gene transfer and transformation. One investigated the effect of exposure to antidepressants on the ability of bacteria to transfer DNA plasmids conferring antibiotic resistance between bacteria by conjugative transfer (20). Another investigated whether antidepressants can accelerate the spread of antibiotic resistance by promoting the transformation of antibiotic resistance genes (ARGs) (26).

### Main Findings

All the included studies established a link between exposure to antidepressants which are in clinical use and promotion of clinically relevant antibiotic resistance in bacteria. Despite the fact that all studies explored different mechanisms of action of antidepressants on antimicrobial resistance, each study identified a positive association between antidepressants and antimicrobial resistance.

An increase in resistance after exposure to antidepressants was confirmed in all of the primary studies (n=8). For example, Jin *et al.,* (2018) established that exposure of *E. coli* to fluoxetine, at a dose that is used clinically in patients with depression, increased the number of bacteria that were resistant to B-lactams, fluoroquinolone, aminoglycosides, chloramphenicol, and tetracycline through ROS mutagenesis (23). Wang *et al.,* (2023) confirmed that antidepressants sertraline, duloxetine, bupropion, escitalopram, and agomelatine at prescription dosage were capable of inducing resistance to multiple antibiotics after brief exposure (30). Li *et al.* (2023) reported that exposing *E. coli* to duloxetine, fluoxetine, amitriptyline, and sertraline led to the development of chloramphenicol-resistant mutants that also exhibited resistance to commonly used environmental disinfectants (25). Shi *et al.,* (2023) analysed the combined effect of antibiotics and antidepressants when used together by monitoring the number, rate of change, and growth pattern of resistant *E. coli* when dosage of chloramphenicol and duloxetine were increased (31).

Several studies reported the dose-response pattern promoting mutation thereby increasing multiple resistance in bacteria. For example, Jin et al. (2018) reported that exposing *E. coli* to fluoxetine at concentrations of 0.5, 5, 50, and 100 mg/L for 30 days significantly increased the mutation frequency (p < 0.01), whereas the lowest concentration of 0.5 mg/L did not result in any increase in mutation (23). Gupinar *et al.,* 2022 also stated that exposure of *A. baumanni* to fluoxetine for 30 days caused a considerable increase in resistance to tetracycline, amoxicillin, and chloramphenicol as well as mutants exhibiting multidrug resistance to different classes of antibiotics (21). Shi *et al.,* 2023 also showed that with an increase in exposure time from a day to 30 days of duloxetine increased the mutation frequency in *E. coli* (13). The two reviews studies emphasize the need for in vivo and population-based studies to explore the link between antidepressants and antimicrobial resistance, suggesting that illness, treatment, or both may influence resistance and increase susceptibility to infections (14, 22).

## DISCUSSION

This scoping review has highlighted associations between antidepressant exposure and clinically relevant antibiotic resistance in laboratory studies. This has potential importance for population health given rising antidepressant prescribing and antibiotic resistance, both global public health crises (6, 8).

### Main findings

The included studies consider different antidepressants from various classes. Fluoxetine and sertraline, the most commonly examined antidepressants in this review, belong to the Selective Serotonin Reuptake Inhibitor (SSRI) class. SSRIs are commonly used in clinical practice, with global prescribing trends reflecting their widespread use across different age groups. According to WHO reports and national surveillance data from countries such as the United States, the United Kingdom, Australia, and Canada, SSRIs consistently rank as the most commonly prescribed antidepressants, regardless of patient age (32–34). SSRIs reduce the mechanism of action of other drugs such as antibiotics (35). At low concentrations sertraline, duloxetine, bupropion, escitalopram, and agomelatine induced antibiotic resistance and persistence in one included study (27). Across all included studies, overall, a positive association between antidepressants exposure and antimicrobial resistance was observed. In some studies, a direct link was observed such as the stimulation of multiple antibiotic resistances in bacteria through ROS-mediated mutagenesis, while in other cases, the role of antimicrobial resistance gene stimulated by some antidepressants was examined. Reactive oxidative stress (ROS) is a significant stress encountered by bacteria as a result of damage to the bacterial cell, antidepressants were confirmed in one study to be responsible for inducing ROS mutagens that are resistant to antibiotics (30). One of the included study utilized PCR and WGS to investigate how 12 different antidepressants influenced antibiotic resistance in *E. coli* and its susceptibility to commonly used disinfectants (25). The study explains that the coexistence of antidepressants, disinfectants and antibiotics in wastewater system may serve as hotspots for the evolution and spread of multidrug resistant bacteria.

### What is already known

Antidepressant drugs have several side effects that can contribute to infection risk in patients, in addition to increasing antibiotic resistance (24). For instance, antidepressants have been associated with increased gastrointestinal infection risk (38), and pneumonia among elderly patients with Parkinson’s disease (36). The use of specific antidepressants has also been associated with the development of *Clostridioides difficile* infections in adults (37). This indicates that beneficial effects of antidepressants, increasing the release of neurotransmitters in the brain improving mood and emotion (38), needs to be balanced with possible negative impact (39).

The most commonly tested antibiotics, for the determination of resistance induced by antidepressants belong to the Beta-lactam class (8). They are the most commonly prescribed antibiotic drugs in the USA and other countries for the treatment of mild to life-threatening infections (8). Clinically, antimicrobial resistance can lead to prolonged, and more severe, illness and increased deaths, globally and can cause significant economic, social, and political disruption (40). The need for population-based studies to determine if associations exist in clinical infections is paramount to improve antidepressant prescribing and reduce antibiotic resistance. Associations observed at the clinical level would have important implications for policy and practice. For instance, these might lead to updates in guidance for individuals on antidepressants who need antibiotics, ensuring the most appropriate antibiotic is prescribed. Additionally, these findings could emphasize the importance of minimizing unnecessary antibiotic use in people on antidepressants to lower their risk of developing antibiotic-resistant infections. They also highlight the value of strengthening non-pharmacological approaches to managing depression, particularly among individuals with recurrent infections.

The first included review highlighted the importance of conducting in vivo and population-based research to determine the association between antidepressants and antimicrobial resistance. It noted that antimicrobial resistance in patients might be influenced by the illness, the treatment, or a combination of both. Additionally, the study suggested that data from patients undergoing antidepressant treatment could help assess whether these individuals face an increased risk of other infections or diseases (22). The second included review summarises emerging evidence that bacteria exposed to antidepressants can develop resistance to multiple antibiotics. It recognises that studies have looked at the mechanisms driving bacterial resistance, and interactions between drugs and the microbes, and highlights the importance of generating evidence to assess the real-world impact of antidepressants on resistance. It also reports that unpublished work examining the microbiota of mice given antidepressants has early data suggesting that antidepressants can alter the gut microbiota and promote gene transfer (14).

### What this review adds

This scoping review maps and synthesizes existing evidence on associations between antidepressants and antimicrobial resistance, including mechanisms involved, and identifies important gaps in literature. There is a notable dearth of evidence concerning the use of antidepressants and their association with antimicrobial resistance, with only 10 studies identified that addressed the review objectives. Existing research has largely been conducted in Australia and China with other regions, with high rates of antidepressant use, underrepresented.

This review identified gaps pertaining to the organisms linked with antimicrobial resistance in patients with depressive disorders. The included studies examined only four bacteria; *E. coli* k12, wild type *E. coli*, *Actinobacter baumani* and *Actinobacter bayigyl* amidst the large numbers of bacteria associated with infections in humans. Different doses, length of exposure, and concentrations of antidepressants and antibiotics were reported across the included studies in this review. Although some studies reported using doses compatible with clinically relevant dosage, direct correlation with potential impact on the gut microbiota of patients cannot be assumed. Therefore, research is required to determine the association between varying doses of antidepressants in humans and their effect on the microbiota, particularly among patients with depressive disorders.

The eight primary research studies included were all laboratory-based, bacterial studies. All found that exposure of bacteria to antidepressant drugs increased antibiotic resistance. The included studies employed a diverse range of laboratory methods to explore the link between antidepressants and antimicrobial resistance. The use of phenotypic and /or genotypic methods in the laboratory to study associations between antidepressants and antimicrobial resistance is an essential step that could provide data to guide treatment decisions. However, laboratory or culture-based investigation can only be applied to external bacteria and is not sufficient for studying human microbiota, as it cannot capture the complexity of the human body (41). The findings from this review support the conclusions of the included review articles which call for research to establish association and to quantify the contribution of non-antibiotic pharmaceuticals to antibiotic resistance (14). In vivo (animal) experiments, the results of which can more readily be applied to human populations, and population studies involving clinical infections, are warranted to determine the impact of associations observed in the laboratory in biological systems. Although, the included reviews addressed antibiotic (antibacterial) resistance only, the search strategy was designed to also retrieve literature relating to antidepressants and, antifungal, antiparasitic, or antiviral resistance. No such publications were retrieved. Similarly, there were no eligible human or animal microbiome studies, or human clinical or population studies.

### Limitations of this review

This is the first review, as far as we know, synthesising existing evidence on the association between antidepressants and antimicrobial resistance. The strengths of this scoping review include, the use of the five-stage approach described by Arksey and O’Malley (15), guided by the PRISMA-ScR checklist, a broad search strategy allowing all study designs to be considered, and a comprehensive overview of the existing evidence. Limiting the included literature to only those written in English language may have resulted in the omission of some eligible studies.

## CONCLUSION

Evidence synthesised in this review identified a links between antidepressants and antibiotic resistance in isolated bacteria across all included studies. No in vivo or population-based studies were retrieved, highlighting the need for more human/animal microbiome studies and clinical/population studies to enable a better understanding of the real-world impact of antidepressants on antimicrobial resistance.

## Author contributions

K.N.B conceptualized the review. B.G.A. was the first reviewer, conducted the literature searches and analysis, and drafted the original manuscript. K.N.B supervised B.G.A. and provided the role of second reviewer. All authors (B.G.A., C.A.M., H.W. & K.N.B,) contributed important intellectual content and revised and edited the manuscript. All authors approved the final manuscript for publication.

## Data Availability

The data underlying this article are available in the article and in its online supplementary material.

## Transparency Declaration

Conflicts of Interest: None to declare.

Funding: There was no funding for this study. An earlier version formed BGA’s Masters dissertation with other authors’ contributions carried out as part of routine work.

## Ethics Statements

Ethical approval is not required for this study as it involves the analysis of publicly available data. Patient consent for publication: Not applicable.

## Notes

### Competing Interest Statement

The authors have declared no competing interest.

