## Supplementary Information for "Association between antidepressants and antimicrobial resistance: a scoping review"

**Supplementary Table I: Search Strategy generated using electronic data base- MEDLINE**

| | **MESH TERM** | **RESULT** |
|---|---|---|
| 1 | ANTIDEPRESSANTS | 83,888 |
| 2 | ANTIDEPRESSIVE DRUGS | 635 |
| 3 | ANTIDEPRESSIVE MEDICATION | 206 |
| 4 | 1 OR 2 OR 3 | 84,296 |
| 5 | ANTIMICROBIAL RESISTANCE | 65,535 |
| 6 | ANTIBACTERIAL RESISTANCE | 3211 |
| 7 | ANTIFUNGAL RESISTANCE | 4084 |
| 8 | ANTIVIRAL RESISTANCE | 3141 |
| 9 | ANTIPROTOZOAL RESISTANCE | 16 |
| 10 | ANTIBIOTIC RESISTANCE | 97,575 |
| 11 | 5 OR 6 OR 7 OR 8 OR 9 OR 10 | 139,431 |
| 12 | 4 AND 11 | 51 |
| 13 | 12 AND (FILTERS – FULL TEXT, ENGLISH, (2013 -2024) YEARS | 57 |

**Supplementary Table II: Search Strategy generated using electronic data base- PUBMED**

| | **MESH TERM** | **RESULT** |
|---|---|---|
| 1 | ANTIDEPRESSANTS | 201,130 |
| 2 | ANTIDEPRESSIVE DRUGS | 44,127 |
| 3 | ANTIDEPRESSIVE MEDICATION | 64,752 |
| 4 | 1 OR 2 OR 3 | 201,130 |
| 5 | ANTIMICROBIAL RESISTANCE | 254,008 |
| 6 | ANTIBACTERIAL RESISTANCE | 226,334 |
| 7 | ANTIFUNGAL RESISTANCE | 34,223 |
| 8 | ANTIVIRAL RESISTANCE | 57,723 |
| 9 | ANTIPROTOZOAL RESISTANCE | 33,185 |
| 10 | ANTIBIOTIC RESISTANCE | 257,211 |
| 11 | 5 OR 6 OR 7 OR 8 OR 9 OR 10 | 414,544 |
| 12 | 4 AND 11 | 822 |
| 13 | 12 AND FILTERS (FULL TEXT, ENGLISH, 10 YEARS | 323 |

Scopus literature search is built to allow selection of subject area, which allows the retrieval of relevant journals within a short time. The broad subject area which includes pharmacology, pharmaceutics, public health, microbiology, and medicine was selected with the search strategy using key words: ANTIDEPRESSANTS OR ANTIDEPRESSIVE AGENTS OR ANTIDEPRESSIVE MEDICATION AND ANTIMICROBIAL RESISTANCE OR ANTIBIOTICS RESISTANCE OR ANTIBACTERIAL RESISTANCE OR ANTIVIRAL RESISTANCE OR ANTIFUNGAL RESISTANCE OR ANTIMICROBIAL RESISTANCE was used to retrieve one hundred and two (102) journals.

For grey literature search, keywords such as “Antidepressants”, “Antimicrobial resistance” and “Antidepressive drugs” was used to retrieve relevant studies.

**Supplementary Table III: List of Excluded Journals.**

| Author/Year | Title | Reason for Exclusion |
|---|---|---|
| Jin et al., 2016 | A Designed Tryptophan- and Lysine/Arginine-Rich Antimicrobial Peptide with Therapeutic Potential for Clinical Antibiotic-Resistant Candida albicans Vaginitis | Not relevant to antidepressants and antimicrobial resistance |
| Dinan and Dinan, 2022 | Antibiotics and mental health: The good, the bad and the ugly | Not relevant to antidepressants and antimicrobial resistance |
| Barnes et al., 2016 | Antidepressant Controlled Trial For Negative Symptoms In Schizophrenia (ACTIONS): a double-blind, placebo-controlled, randomised clinical trial | Not relevant to antidepressants and antimicrobial resistance |
| Deslouches et al., 2016 | Comparative functional properties of engineered cationic antimicrobial peptides consisting exclusively of tryptophan and either lysine or arginine | Not relevant to antidepressants and antimicrobial resistance |
| Martins et al., 2020 | Comparing activity, toxicity and model membrane interactions of Jelleine-I and Trp/Arg analogs: analysis of peptide aggregation | Not relevant to antidepressants and antimicrobial resistance |
| Howan et al., 2023 | Enhanced Antibacterial Activity of Substituted Derivatives of NCR169C Peptide | Not relevant to antidepressants and antimicrobial resistance |
| Blanco-Rivero et al., 2021 | Enhanced sympathetic neurotransduction in the superior mesenteric artery in a rat model of heart failure: role of noradrenaline and ATP | Not relevant to antidepressants and antimicrobial resistance |
| Frontana et al., 2020 | Exploring the role of gut microbiota in major depressive disorder and in treatment resistance to antidepressants | Not relevant to antidepressants and antimicrobial resistance |
| Dong et al., 2022 | Gut microbiome: A potential indicator for predicting treatment outcomes in major depressive disorder | Not relevant to antidepressants and antimicrobial resistance |
| Xu et al., 2023 | Interactions Between Antidepressants and Intestinal Microbiota | Not relevant to antidepressants and antimicrobial resistance |
| Chen et al., 2019 | Maintenance of antidepressant and anti-suicidal effects by D-cycloserine among patients with treatment-resistant depression who responded to low-dose ketamine infusion: a double-blind randomized placebo-control study | Not relevant to antidepressants and antimicrobial resistance |

| Author/Year | Title | Reasons for exclusion |
|---|---|---|
| Ronaldson et al., 2022 | Prospective associations between depression and risk of hospitalisation for infection: Findings from the UK Biobank | Not relevant to antidepressants and antimicrobial resistance |
| Arias et al., 2016 | Recombinant expression, antimicrobial activity and mechanism of action of tritrpticin analogs containing fluoro-tryptophan residues | Not relevant to antidepressants and antimicrobial resistance |
| Kopoor et al., 2020 | Repurposing of Existing Drugs for the Bacterial Infections: An In silico and In vitro Study | Not relevant to antidepressants and antimicrobial resistance |
| Batista et al., 2019 | A mechanistic approach to the in-vitro resistance modulating effects of fluoxetine against meticillin resistant Staphylococcus aureus strains | Not relevant to antidepressants and antimicrobial resistance |
| Wilson et al., 2018 | Resensitization of methicillin-resistant Staphylococcus aureus by amoxapine, an FDA-approved antidepressant | Not relevant to antidepressants and antimicrobial resistance |
| Meisel et al., 2016 | Reversal of Tetracycline Resistance in Escherichia coli by Noncytotoxic bis (Tryptophan)s | Not relevant to antidepressants and antimicrobial resistance |
| Chaudhary et al., 2017 | Safety and efficacy of a novel drug elores (ceftriaxone + sulbactam + disodium edetate) in the management of multi-drug resistant bacterial infections in tertiary care centers: a post-marketing surveillance study | Not relevant to antidepressants and antimicrobial resistance |
| Liu et al., 2019 | Synthesis and biological evaluation of tryptophan-derived rhodanine derivatives as PTP1B inhibitors and anti-bacterial agents | Not relevant to antidepressants and antimicrobial resistance |
| Seeman 2021 | The gut microbiome and antipsychotic treatment response | Not relevant to antidepressants and antimicrobial resistance |
| Kromann et al., 2017 | Treatment with high-dose antidepressants severely exacerbates the pathological outcome of experimental Escherichia coli infections in poultry | Not relevant to antidepressants and antimicrobial resistance |
| Ou J 2023 | Unintended Microbial Targets of Antidepressants | Not relevant to antidepressants and antimicrobial resistance |
| Osborn et al., 2013 | Antidepressant-like effects of erythropoietin: a focus on behavioural and hippocampal processes | Not relevant to antidepressants and antimicrobial resistance |

| Author/Year | Title | Reason for Exclusion |
|---|---|---|
| Crane et al., 2021 | Psychoactive Drugs Induce the SOS Response and Shiga Toxin Production in Escherichia coli | Focused on antimicrobial resistance of Psychotic drugs which is not relevant to the scope of this research |
| Ellezian et al., 2022 | Psychotropic Drugs in the Discussion of Antimicrobial-Resistant Microorganisms | Focused on antimicrobial resistance of Psychotic drugs which is not relevant to the scope of this research |
| Kyono et al., 2021 | The Atypical Antipsychotic Quetiapine Promotes Multiple Antibiotic Resistance in Escherichia coli | Focused on antimicrobial resistance of Psychotic drugs which is not relevant to the scope of this research |
| Cussotto et al., 2021 | Psychotic drugs and the Microbes | Focused on antimicrobial resistance of Psychotic drugs which is not relevant to the scope of this research |
| Bachtarzi et al., 2019 | Psychoactive drug prescription and urine colonization with extended-spectrum β-lactamase-producing enterobacteriaceae | Focused on antimicrobial resistance of Psychotic drugs which is not relevant to the scope of this research |
| Oliveira et al., 2018 | Anti-Candida activity of antidepressants sertraline and fluoxetine: effect upon pre-formed biofilms | Focused on antimicrobial activities of antidepressants |
| Ahmed et al., 2023 | Anti-Candidal Activity of Reboxetine and Sertraline Antidepressants: Effects on Pre-Formed Biofilms | Focused on antimicrobial activities of antidepressants |
| Bottega et al., 2020 | Antimicrobial and Antineoplastic Properties of Sertraline | Focused on antimicrobial activities of antidepressants |
| Dos Santos et al., 2020 | Antibacterial activity of fluoxetine-loaded starch nanocapsules | Focused on antimicrobial activities of antidepressants |
| Gonzalez et al., 2022 | Antidepressants with antimicrobial activity? A promising therapeutic strategy | Focused on antimicrobial activities of antidepressants |
| Macedo et al., 2017 | Antidepressants, antimicrobials or both? | Focused on antimicrobial activities of antidepressants |
| Macedo et al., 2017 | Gut microbiota dysbiosis in depression and possible implications of the antimicrobial effects of antidepressant drugs for antidepressant effectiveness | Focused on antimicrobial activities of antidepressants |

| **Author/Year** | **Title** | **Reason for Exclusion** |
|---|---|---|
| Caldara and Marmiroli 2021 | Antimicrobial properties of antidepressants and antipsychotics—Possibilities and implications | Focused on antimicrobial activities of antidepressants |
| Lagadinou et al., 2020 | Antimicrobial properties on non-antibiotic drugs in the era of increased bacterial resistance | Focused on antimicrobial activities of antidepressants |
| Andersson et al., 2018 | Combating Multidrug-Resistant Pathogens with Host-Directed Nonantibiotic Therapeutics | Focused on antimicrobial activities of antidepressants |
| Mohammed et al., 2015 | Citalopram and venlafaxine differentially augments Antimicrobial properties of antibiotics | Focused on antimicrobial activities of antidepressants |
| Shoaib et al., 2023 | Evaluating Fluoxetine's Growth-Inhibitory Impact on Clinical Isolates of Enteric Bacteria: Harnessing Repurposing Potential | Focused on antimicrobial activities of antidepressants |
| Josino et al., 2021 | Development and in vitro evaluation of microparticles of fluoxetine in galactomannan against biofilms of S. aureus methicilin resistant | Focused on antimicrobial activities of antidepressants |
| Hussein et al., 2020 | Effective Strategy Targeting Polymyxin-Resistant Gram-Negative Pathogens: Polymyxin B in Combination with the Selective Serotonin Reuptake Inhibitor Sertraline | Focused on antimicrobial activities of antidepressants |
| Alidjinou et al., 2019 | Emergence of Fluoxetine-Resistant Variants during Treatment of Human Pancreatic Cell Cultures Persistently Infected with Coxsackievirus B4 | Focused on antimicrobial activities of antidepressants |
| Villanueva-Lozano et al., 2020 | Evaluation of the expanding spectrum of sertraline against uncommon fungal pathogens. | Focused on antimicrobial activities of antidepressants |
| Bauer et al., 2019 | Fluoxetine Inhibits Enterovirus Replication by Targeting the Viral 2C Protein in a Stereospecific Manner | Focused on antimicrobial activities of antidepressants |
| Lieb J, 2004 | The immunostimulating and antimicrobial properties of lithium and antidepressants | Focused on antimicrobial activities of antidepressants |
| Krzyzek et al., 2019 | In Vitro Activity of Sertraline, an Antidepressant, Against Antibiotic-Susceptible and Antibiotic-Resistant Helicobacter pylori Strains | Focused on antimicrobial activities of antidepressants |

| Author/Year | Title | Reasons for Exclusion |
|---|---|---|
| Silva et al., 2017 | In vitro anti-Candida activity of selective serotonin reuptake inhibitors against fluconazole-resistant strains and their activity against biofilm-forming isolates | Focused on antimicrobial activities of antidepressants |
| Cong et al., 2016 | In Vitro Antifungal Activity of Sertraline and Synergistic Effects in Combination with Antifungal Drugs against Planktonic Forms and Biofilms of Clinical Trichosporon asahii Isolates | Focused on antimicrobial activities of antidepressants |
| Li et al., 2017 | Insight into synergetic mechanisms of tetracycline and the selective serotonin reuptake inhibitor, sertraline, in a tetracycline-resistant strain of Escherichia coli | Focused on antimicrobial activities of antidepressants |
| De Sousa et al., 2018 | New roles of fluoxetine in pharmacology: Antibacterial effect and modulation of antibiotic activity | Focused on antimicrobial activities of antidepressants |
| Barbarossa et al., 2022 | Non-Antibiotic drug repositioning as an alternative antimicrobial approach | Focused on antimicrobial activities of antidepressants |
| Laudy 2018 | Non-antibiotics, Efflux Pumps and Drug Resistance of Gram-negative Rods | Focused on antimicrobial activities of antidepressants |
| Foletto et al., 2021 | Repositioning of antidepressant drugs and synergistic effect with ciprofloxacin against multidrug-resistant bacteria | Focused on antimicrobial activities of antidepressants |
| Da Rosa et al., 2020 | Repositioning or redirection of antidepressant drugs in the treatment of bacterial and fungal infections | Focused on antimicrobial activities of antidepressants |
| Racz and Spengler, 2023 | Repurposing Antidepressants and Phenothiazine Antipsychotics as Efflux Pump Inhibitors in Cancer and Infectious Diseases | Focused on antimicrobial activities of antidepressants |
| Rosa et al., 2021 | Repurposing of escitalopram oxalate and clonazepam in combination with ciprofloxacin and sulfamethoxazole-trimethoprim for treatment of multidrug-resistant microorganisms and evaluation of the cleavage capacity of plasmid DNA | Focused on antimicrobial activities of antidepressants |

| Author/Year | Title | Reason for Exclusion |
|---|---|---|
| Geeraerts et al., 2021 | Repurposing the antidepressant sertraline as SHMT inhibitor to suppress serine/glycine synthesis–addicted breast tumor growth. | Focused on antimicrobial activities of antidepressants |
| McGovern et al., 2019 | A review of the antimicrobial side of antidepressants and its putative implications on the gut microbiome | Focused on antimicrobial activities of antidepressants |
| Kubra YILDIRIM, 2023 | Investigation of the Antituberculosis Effectiveness of Escitalopram, a Selective Serotonin Reuptake Inhibitor | Focused on antimicrobial activities of antidepressants |
| Zinnah and Park, 2021 | Sensitizing TRAIL-resistant A549 lung cancer cells and enhancing TRAIL-induced apoptosis with the antidepressant amitriptyline | Focused on antimicrobial activities of antidepressants |
| Alkhalifa et al., 2022 | Serotonin reuptake inhibitors effect on fluconazole activity against resistant Candida glabrata strains | Focused on antimicrobial activities of antidepressants |
| Ayaz et al., 2015 | Sertraline enhances the activity of antimicrobial agents against pathogens of clinical relevance | Focused on antimicrobial activities of antidepressants |
| Rodrigues et val., 2023 | Sertraline has in vitro activity against both mature and forming biofilms of different Candida species | Focused on antimicrobial activities of antidepressants |
| Rocha et al., 2022 | Synergism between the Antidepressant Sertraline and Caspofungin as an Approach to Minimise the Virulence and Resistance in the Dermatophyte Trichophyton rubrum | Focused on antimicrobial activities of antidepressants |
| Ou et al., 2022 | TCA and SSRI Antidepressants Exert Selection Pressure for Efflux-Dependent Antibiotic Resistance Mechanisms in Escherichia coli | Focused on antimicrobial activities of antidepressants |
| Da Silva Rodrigues et al., 2019 | The antidepressant clomipramine induces programmed cell death in Leishmania amazonensis through a mitochondrial pathway | Focused on antimicrobial activities of antidepressants |
| Breuer et al., 2022 | The Antidepressant Sertraline Induces the Formation of Supersized Lipid Droplets in the Human Pathogen Cryptococcus neoformans | Focused on antimicrobial activities of antidepressants |
| Shankaran et al., 2023 | The antidepressant sertraline provides a novel host directed therapy module for augmenting TB therapy | Focused on antimicrobial activities of antidepressants |

| **Author/Year** | **Title** | **Reason for Exclusion** |
|---|---|---|
| Gu et al., 2023 | The Synergistic Effect of Azoles and Fluoxetine against Resistant Candida albicans Strains Is Attributed to Attenuating Fungal Virulence | Focused on antimicrobial activities of antidepressants |
| Gillard et al., 2018 | Tricyclic amine antidepressants suppress β-lactam resistance in methicillin-resistant Staphylococcus aureus (MRSA) by repressing mRNA levels of key resistance genes | Focused on antimicrobial activities of antidepressants |
| Caldara and Marmiroli 2018 | Tricyclic antidepressants inhibit Candida albicans growth and biofilm formation | Focused on antimicrobial activities of antidepressants |
| Ait Chait et al., 2022 | Unravelling the antimicrobial action of antidepressants on gut commensal microbes | Focused on antimicrobial activities of antidepressants |

**Supplementary Table IV: Quality assessment of included studies**

| **Author / Year** | **Score** | **Rating** |
|---|---|---|
| Drew L. (2023) (14) | 3 | Not assignable |
| Hategan and Bourgeois (2023) (22) | 3 | Not assignable |
| Lavier L (2023) (24) | 12 | Reliable with restrictions |
| Gurpinar *et al.* (2022) (21) | 12 | Reliable with restrictions |
| Jin *et al.* (2018) (23) | 12 | Reliable with restrictions |
| Shi *et al.* (2022) (13) | 15 | Reliable without restrictions |
| Ding *et al.* (2022) (20) | 16 | Reliable without restrictions |
| Lu *et al.* (2022) (26) | 16 | Reliable without restrictions |
| Wang *et al.* (2023) (27) | 16 | Reliable without restrictions |
| Li *et al.* (2023) (29) | 17 | Reliable without restrictions |
